# “Somebody to find an answer, so I can sleep at night”: Diverse Perspectives on Genomic Research for Rare Disease

**DOI:** 10.64898/2026.10.23.26366105

**Authors:** Ashana Neale, Jacklyn O. Durigan, Hadley Stevens Smith, Yarden S. Fraiman, Siwaar Abouhala, Eva Martinez, Heidi L. Rehm, Anne O’Donnell-Luria, Melanie C. O’Leary, Monica H. Wojcik

**Affiliations:** Center for Mendelian Genomics, Program in Medical and Population Genetics, Broad Institute of MIT and Harvard, Cambridge, Massachusetts, USA; BCH; Department of Neonatoloy, Beth Israel Deaconess Medical Center, Boston, MA. Harvard Medical School, Boston, MA; Center for Genomic Medicine, Massachusetts General Hospital, Boston, MA, USA. Center for Mendelian Genomics, Program in Medical and Population Genetics, Broad Institute of MIT and Harvard, Cambridge, MA, USA. Department of Pathology, Harvard Medical Schoo; Center for Genomic Medicine, Massachusetts General Hospital, Boston, MA, USA. Program in Medical and Population Genetics, Broad Institute of MIT and Harvard, Cambridge, MA, USA., Division of Genetics and Genomics, Boston Children’s Hospital, Harvard Medic; Center for Mendelian Genomics, Program in Medical and Population Genetics, Broad Institute of MIT and Harvard, Cambridge, Massachusetts, USA; Divisions of Newborn Medicine and of Genetics and Genomics, Boston Children’s Hospital, Harvard Medical School, B

## Abstract

**Purpose:** Rare disease genomic research suffers from the underrepresentation of minoritized populations. We therefore sought to elucidate equitable strategies for participant engagement in rare disease genomic research.

**Methods:** Eligible participants had previously applied to the Rare Genomes Project (RGP) and are underrepresented in genomic research by virtue of race, ethnicity, language, socio-economic status, educational level, or rural status. We also recruited clinicians who have referred potential participants to RGP. We performed semi-structured interviews with all enrollees.

**Results:** Analysis of the 41 interviews—including 25 RGP participants and 16 clinicians—yielded four primary themes: 1) rare disease genomic research studies are intertwined with clinical care and hold potential to overcome access barriers in the clinic, with subsequent equity implications, 2) research participation functions as a rare disease community resource, offering psychosocial benefit and access to emerging science, 3) flexibility in research processes allows for increased access for diverse participants, and 4) longitudinal engagement is strengthened through relational continuity with participants and their clinical team.

**Conclusion:** Overall, the motivators and experiences shared by clinicians and diverse participants inform effective engagement in rare disease genomic research and highlight the importance of providing equitable access to genomic sequencing for rare disease diagnosis.

## Introduction

Rare diseases affect up to 10% of the global population^1, 2^ and have a disproportionate impact on public health, representing the leading cause of infant mortality^3, 4^ and comprising nearly one-half of pediatric healthcare expenditures in the United States^5, 6^. Identifying a molecular diagnosis for individuals with rare diseases can be life-changing by ending the diagnostic odyssey, facilitating better prognostication, guiding surveillance, and allowing for precision therapies, when available^7^. Recent technological innovations, such as high-throughput pipelines for genome sequencing (GS), have expanded the ability to detect genomic variants responsible for disease^8^. The capacity for sequencing, however, has advanced at a faster rate than our understanding of the genomic data produced, and interpretation of genomic variation may require significant research. For example, the identification of non-coding RNU genes as contributors to syndromic neurodevelopmental disease required large rare disease genomic research consortia^9, 10^. Consequently, genomic research plays a vital role in current applications of genomic medicine for people with rare diseases In addition to advancing discovery of gene-disease relationships and facilitating variant classification and interpretation, research programs also provide an avenue for individuals to access diagnostic testing that may otherwise not be feasible due to insurance denials, cost constraints, or limited clinical availability^11–14^.

However, when considering the benefits of genomic research across communities, important ethical concerns remain. Minoritized populations are underrepresented in rare disease genomic research^15^ and inequities in access to genomic services, both research and clinical, leads to inequitable outcomes^16^. Addressing these disparities requires a deeper understanding of the barriers and facilitators influencing enrollment in genomic research for potential participants.

The Rare Genomes Project (RGP) is a rare disease genomic research study with the goal of discovering the underpinnings of rare Mendelian diseases via direct engagement with self-referred, genetically undiagnosed patients via remote interactions with the RGP team, bypassing the need to travel to a study site^17^. In a prior pilot study^17^, we evaluated the acceptability and adoption of enhanced community engagement and supportive enrollment for populations historically underrepresented in genomic research. We were able to support diverse representation within RGP by implementing online clinician referral forms, non-English language support, off-hours staff availability for participant engagement, and mobile phlebotomy services for sample collection^18^. We now aim to further understand experiences of clinicians and individuals from minoritized populations with rare disease in applying to and participating in RGP to support inclusive and equitable recruitment both in this study and ultimately other genomic research or even clinical initiatives.

## Methods

The Rare Genomes Project was launched in 2017 at the Broad Institute of MIT and Harvard and offers research GS for individuals and families with rare, genetically undiagnosed conditions. As a research study, participation is free of cost. Eligibility criteria include having clinical suspicion of a monogenic condition, lacking a confirmed genetic diagnosis, and residing in the United States. Prospective participants may submit an application via the study website or be referred by their clinician. Applications and referral forms are reviewed to determine eligibility. For accepted families, RGP study staff work with families to obtain informed consent remotely (typically via electronic consent forms and telephone or video calls with RGP staff) and coordinate remote collection of blood samples from the affected individual and relevant family members, which are sent to the Broad Institute of MIT and Harvard for GS and analysis. Diagnostic variants are confirmed in a clinical laboratory and returned to the participant, often in collaboration with a clinician of their choice.

### Recruitment

The invitation to participate in this qualitative research study was emailed to families who had previously enrolled in RGP or who had initiated but not completed the enrollment process and who were underrepresented in our study cohort by race (race other than white), ethnicity (Hispanic or Latino ethnicity), primary language (other than English), household income (under federal poverty line), education level (high school or less), or rural residence (defined as Rural-Urban Commuting Area score^19^ greater than or equal to four). Families who did not opt out of future contact following the initial email invitation were also contacted by phone. For clinician participants, email invitations were distributed to clinicians who had previously referred historically underrepresented participants to RGP. Following expression of interest in enrolling in this study via reply to the email invitation or agreement to participate via phone call, interviews were scheduled to occur over video conferencing or telephone call for those unable to access video conferencing technologies.

### Interview Guide Development

The semi-structured interview guides (one for the RGP participants and one for the referring clinicians) were collaboratively developed by our research team to elucidate barriers and facilitators of genomic research participation. These guides were designed to address the domains presented in **Table 1**, informed by the Consolidated Framework for Implementation Research^20^ (CFIR). We invoke the CFIR in this design due to the need to contextualize rare disease genomic research participation within current systems of genomic care, and to generate findings relevant and translatable to clinical practice. Demographic characteristics of interviewees were collected via self-report at the end of the interview. An external advisory board, comprising 5 individuals from historically medically underserved backgrounds (four of whom are personally affected or have a child affected by a rare genetic condition) reviewed the interview guide and provided formative feedback on the questions and content in discussions via video conferences. Final interview guides are provided in the Appendix.

**Table 1.** Domains of semi-structured interview guide for participants and referring clinicians.

| **Cohort** | **Semi-structured interview guide domain** | **CFIR Domain and Construct** |
| --- | --- | --- |
| **Participants**<br>individuals referred to RGP from underserved populations | Sociocultural beliefs and impact on research participation: e.g., trust, cultural values | Outer setting: local attitudes |
|  | Mechanism by which participant heard about the research study and influence on enrollment | Outer setting: partnerships and connections |
|  | Study logistics as a barrier or facilitator | Inner setting: infrastructure |
|  | Interactions with study staff as barriers or facilitators | Inner setting: relationships and connections |
|  | Perceived culture of the study team and impact on participation | Inner setting: culture |
|  | Perceived motivations and desire to participate in genomic research as well as ability to do so | Individual: need, motivation, opportunity capability |
| **Clinicians**<br>clinicians with a history of referral of underserved participants to RGP | Policies as barriers to genomic medicine access and impact on equity | Outer setting: policies |
|  | Impact of clinic resources on research referral | Outer setting: financing |
|  | Perception of shared goals between clinician and researcher | Inner setting: mission alignment |
|  | Study logistics and resources as a barrier or facilitator | Inner setting: resources |
|  | Perceived motivations and desire to participate in genomic research as well as ability to do so | Individual: need, capability, opportunity, motivation |

### Data Collection

Interviews lasted 30-60 minutes and were conducted in English or Spanish by one of three members of the research team (S.A., E.M., A.N.) with a second member present on videoconferences to facilitate recording (A.T.M.). Interviews conducted with Spanish-speaking families were conducted by a bilingual team member (E.M.). Upon completion of each interview, interviewees were compensated for their time with a $50 gift card.

### Analysis

Interview transcripts were professionally transcribed and de-identified. Spanish transcripts were translated to English by the transcription service. The transcripts were pooled and independently coded by three authors in a mixed deductive-inductive approach. Initial deductive codes were developed based on the stages of RGP study participation. Inductively developed codes were discussed among the research team to collaboratively develop a codebook. Using the final codebook, all transcripts were independently coded by three study team members (A.N., M.H.W., J.O.D.) using NVivo. Codes were discussed and organized into inductively developed themes that were iteratively discussed and refined among the coders (A.N., M.H.W., J.O.D.) and with the study team. These same coders, who were a genetic counselor and two medical geneticists, then used the themes to inform recommendations for inclusive rare disease genomic research and clinical care.

## Results

### Participant and Clinician Demographics

We completed interviews with 25 RGP participants (18 in English and 7 in Spanish) and 16 clinicians, for a total of 41 interviews. Demographics of RGP participants and clinicians are presented in **Table 2**.

**Table 2.**
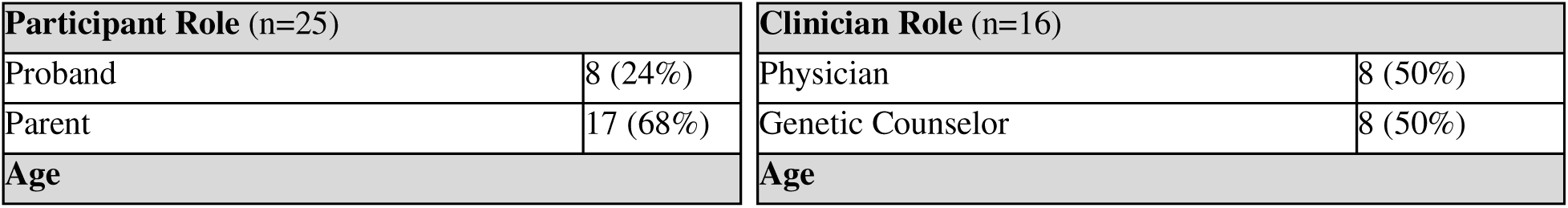
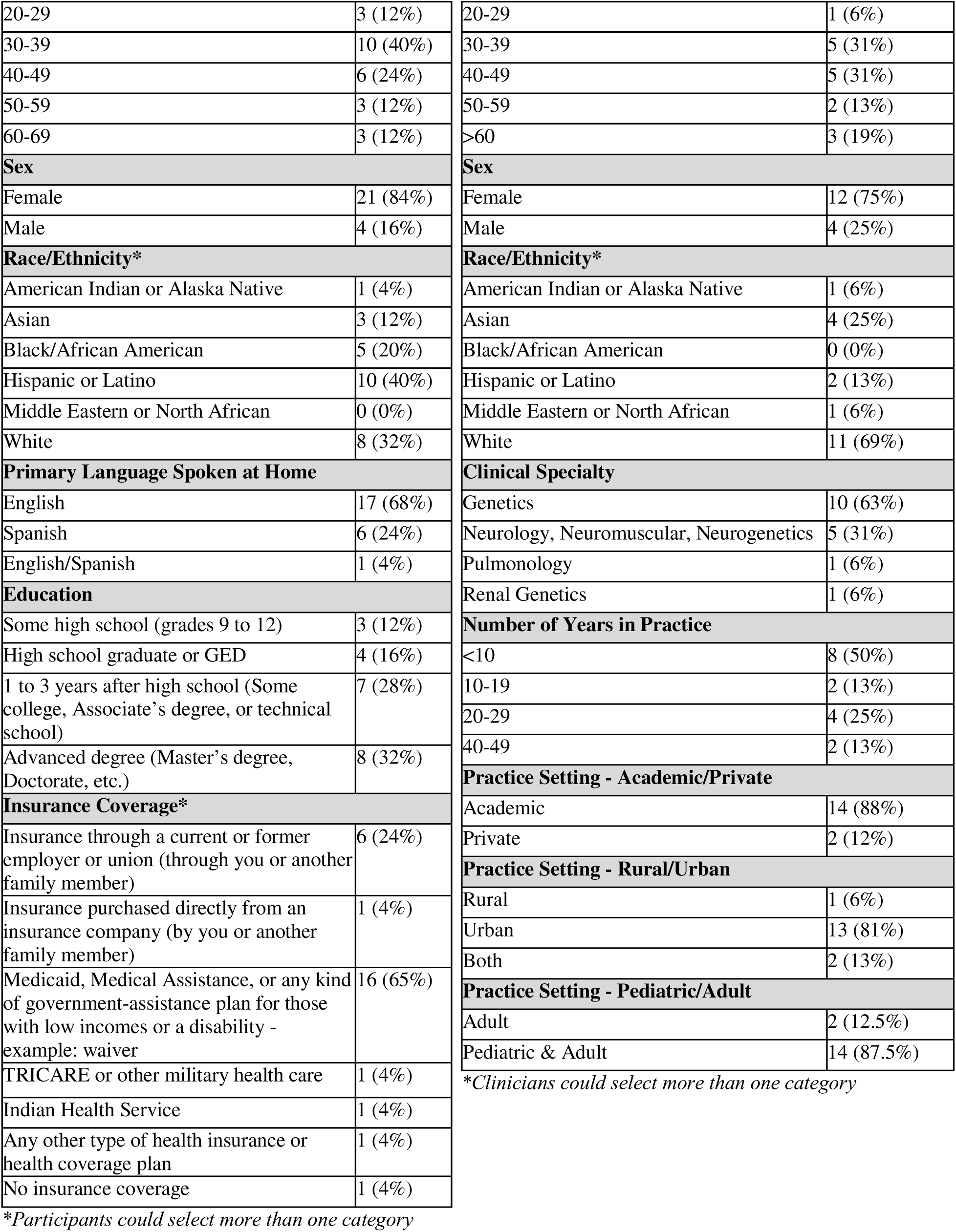
Participant and Clinician Demographic Information.

| Participant Role (n=25) |  | Clinician Role (n=16) |  |
| --- | --- | --- | --- |
| Proband | 8 (24%) | Physician | 8 (50%) |
| Parent | 17 (68%) | Genetic Counselor | 8 (50%) |
| Age |  | Age |  |

|  |  |
| --- | --- |
| 20-29 | 3 (12%) |
| 30-39 | 10 (40%) |
| 40-49 | 6 (24%) |
| 50-59 | 3 (12%) |
| 60-69 | 3 (12%) |
| **Sex** |  |
| Female | 21 (84%) |
| Male | 4 (16%) |
| **Race/Ethnicity\*** |  |
| American Indian or Alaska Native | 1 (4%) |
| Asian | 3 (12%) |
| Black/African American | 5 (20%) |
| Hispanic or Latino | 10 (40%) |
| Middle Eastern or North African | 0 (0%) |
| White | 8 (32%) |
| **Primary Language Spoken at Home** |  |
| English | 17 (68%) |
| Spanish | 6 (24%) |
| English/Spanish | 1 (4%) |
| **Education** |  |
| Some high school (grades 9 to 12) | 3 (12%) |
| High school graduate or GED | 4 (16%) |
| 1 to 3 years after high school (Some college, Associate's degree, or technical school) | 7 (28%) |
| Advanced degree (Master's degree, Doctorate, etc.) | 8 (32%) |
| **Insurance Coverage\*** |  |
| Insurance through a current or former employer or union (through you or another family member) | 6 (24%) |
| Insurance purchased directly from an insurance company (by you or another family member) | 1 (4%) |
| Medicaid, Medical Assistance, or any kind of government-assistance plan for those with low incomes or a disability - example: waiver | 16 (65%) |
| TRICARE or other military health care | 1 (4%) |
| Indian Health Service | 1 (4%) |
| Any other type of health insurance or health coverage plan | 1 (4%) |
| No insurance coverage | 1 (4%) |

\*Participants could select more than one category

|  |  |
| --- | --- |
| 20-29 | 1 (6%) |
| 30-39 | 5 (31%) |
| 40-49 | 5 (31%) |
| 50-59 | 2 (13%) |
| >60 | 3 (19%) |
| **Sex** |  |
| Female | 12 (75%) |
| Male | 4 (25%) |
| **Race/Ethnicity\*** |  |
| American Indian or Alaska Native | 1 (6%) |
| Asian | 4 (25%) |
| Black/African American | 0 (0%) |
| Hispanic or Latino | 2 (13%) |
| Middle Eastern or North African | 1 (6%) |
| White | 11 (69%) |
| **Clinical Specialty** |  |
| Genetics | 10 (63%) |
| Neurology, Neuromuscular, Neurogenetics | 5 (31%) |
| Pulmonology | 1 (6%) |
| Renal Genetics | 1 (6%) |
| **Number of Years in Practice** |  |
| <10 | 8 (50%) |
| 10-19 | 2 (13%) |
| 20-29 | 4 (25%) |
| 40-49 | 2 (13%) |
| **Practice Setting - Academic/Private** |  |
| Academic | 14 (88%) |
| Private | 2 (12%) |
| **Practice Setting - Rural/Urban** |  |
| Rural | 1 (6%) |
| Urban | 13 (81%) |
| Both | 2 (13%) |
| **Practice Setting - Pediatric/Adult** |  |
| Adult | 2 (12.5%) |
| Pediatric & Adult | 14 (87.5%) |

\*Clinicians could select more than one category

Of the 25 RGP participants, most (68%) were parents of a child with a rare genetic disorder and the remainder had a rare disease themselves. The majority of the cohort identified as female (84%). Their reported racial and ethnic backgrounds were diverse, and the seven Spanish-speaking participants (28%) reported using a medical interpreter during healthcare encounters. Additional demographic characteristics are reported in **Table 2**. Most participants had completed all aspects of study participation, although 9/25 failed to complete all study steps, including submitting a sample for data analysis or the necessary medical records to support data analysis. (**Figure 1**).

**Figure 1.**
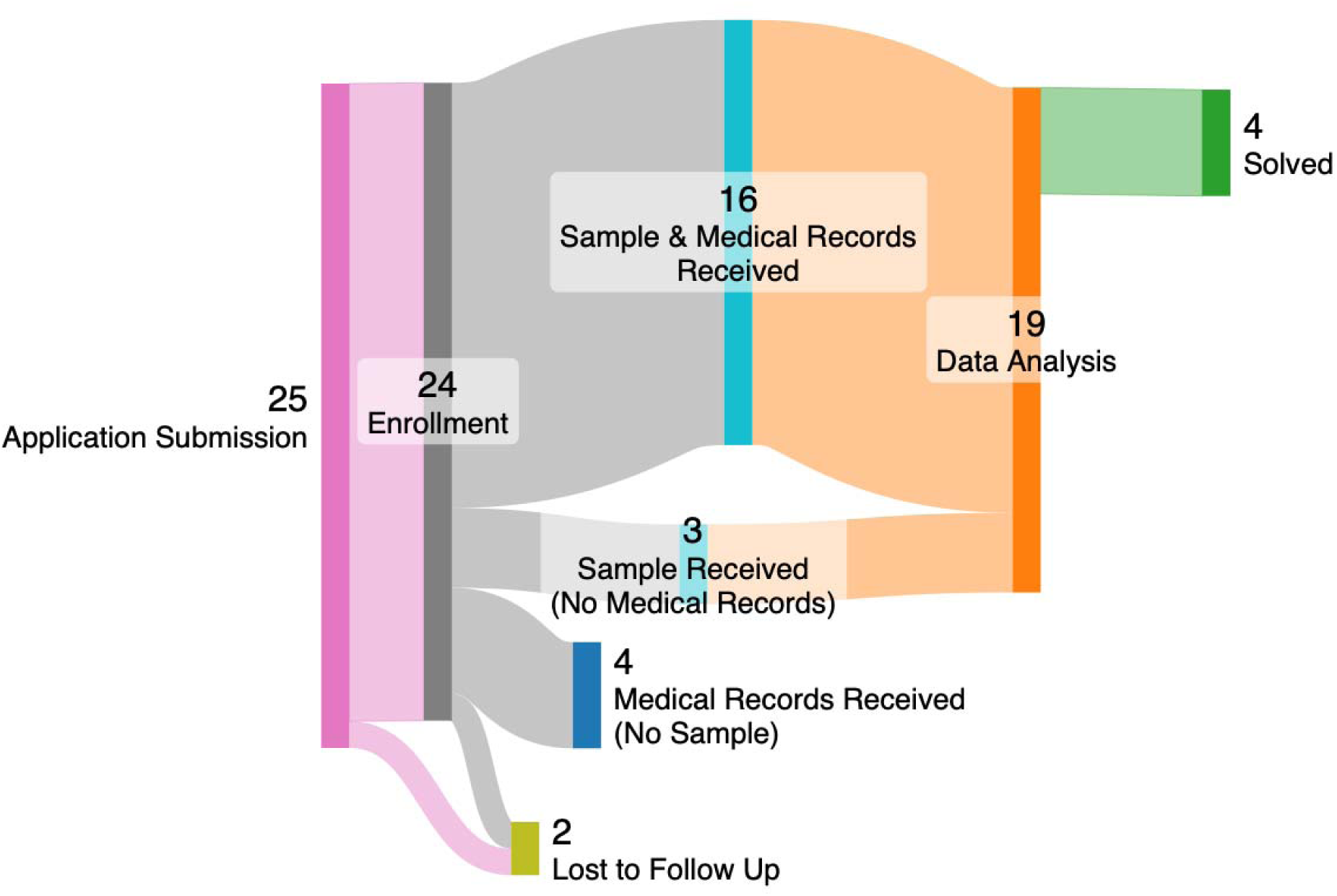
Participant flow through the Rare Genomes Project timeline from initial application submission to result return. The Sankey plot illustrates the 25 participant interviewees progressing through each stage of enrollment and participation, indicating where participants were at the time of their interview. Following confirmation of eligibility after application submission, one participant did not engage in a study consent session and was lost to follow-up. Twenty-four individuals successfully finalized the consent process and proceeded to study enrollment. One additional participant was lost-to-follow-up after enrollment, resulting in no sample acquisition or medical record collection. Four additional participants did not submit their samples for GS but did provide us with relevant medical records. Nineteen participants submitted their samples, with four considered solved after GS analysis and fifteen with genomic data analysis currently ongoing. The Sankey Matic tool was used to obtain the Sankey diagram.

Of the 16 clinicians interviewed, there were an equal number of physicians and genetic counselors. The majority identified as female (75%), and most clinicians identified as White, followed by Asian. While a majority worked within the genetic clinical specialty, other specialties were represented, including neurology and pulmonology. **Table 2** provides other demographic features.

### Thematic Analysis

We developed four primary themes (**Table 4**), which reflect the process and context of genomic medicine and genomic research for families with rare diseases and highlight particular considerations for historically underserved and underrepresented populations. Themes and subthemes are described in detail in the sections that follow.

#### Theme 1: Rare disease genomic research studies are intertwined with clinical care and hold potential overcome access barriers in the clinic, with subsequent equity implications

Participants entered genomic research studies from a background of prior clinical care that at times felt supportive or not supportive of their clinical goals. Both participants and clinicians noted that when available testing options have been exhausted, either due to cost or lack of commercial offerings, research can provide a mechanism to continue the diagnostic odyssey. While genetics clinicians explicitly pointed to their use of rare disease research studies to access genomic technologies that are not available in the clinical setting due to financial barriers, the research participants conceptualized the research setting providing access to new and emerging advances in genomic diagnostics without as much of a transactional view regarding access to a specific test (GS). Subthemes within this overarching theme included 1) perceived clinical utility of genomic research, 2) research to improve the care of others, and 3) research as an extension of clinical care. These subthemes also relate to equitable access to genomic medicine and the ways in which research can either promote or undermine equitable access in rare disease genomics.

#### Subtheme 1: Perceived clinical utility of research

Both clinicians and participants noted that a major motivator to participate in rare disease genomic research is the hope that a result will be identified that may directly impact clinical care. Common desired benefits identified included the ability to know if a condition could be passed down to one’s children and finding possible treatments or improved management strategies for their or their child’s symptoms. Some participants had tangible benefits in mind, such as medication for the treatment of symptoms. Others were hoping for cures that are yet to come, possibly reflecting participation in research as a way to access not only novel diagnostics but also novel therapeutics.

A unique feature of rare genetic conditions is that many are heritable, and thus the potential to help additional family members, notably other children, was another driver of rare disease research participation. In particular, families with one affected child were worried about whether their other children may also develop the same symptoms due to a heritable condition. Others were open to any tangible clinical impact that research participation could have. For example, one participant (Participant 8, Asian male, parent, analysis in progress) described looking for any way to improve their child’s symptoms, not necessarily a “magic wand.” One clinician stated (Clinician 15, Asian female, pediatric genetic counselor) stated that a concrete medical impact may not be the most important factor to parents, when just the knowledge itself may be helpful, if not to “cure” the disease, to “cure” the feeling of being undiagnosed: “Somebody to find an answer, so I can sleep at night.” (Participant 13, White Hispanic female, parent, did not enroll). Indeed, the idea that knowing the diagnosis would open doors to clinical utility, such as management changes or targeted therapies, was commonly stated. For many participants, this potential for a positive clinical impact far outweighed any perceived downside to enrollment. Other tangible benefits included impact on other domains of care, such as finding new medications or improving methods of communication, where one participant expressed a desire to “get some more insight as to, you know, what’s wrong with him, and maybe to even be able to help us to be able to communicate better with him…. I think that would be a wonderful thing.” (Participant 1, Black or African American female, parent, sample pending).

For participants from populations facing barriers to genomic medicine access, the potential for clinical benefit may be more significant than to those not facing historical barriers. One participant noted that their sister “…has several conditions similar to all of this. But the difference is that they haven’t been checked out by a doctor” (Participant 10, Hispanic male, Spanish-speaking proband, analysis in progress). The notion of genomic research participation as an extension of clinical care is deeply intertwined with the financial complexity of access to diagnostic technologies such as genome sequencing. Due to the out-of-pocket cost of genetic testing, insurance coverage is a requirement for access for many people with rare diseases. Indeed, one clinician stated that they do not refer to our research study often “because we do have a lot of patients who are motivated to pay out of pocket,” (Clinician 14, White Hispanic female, neurology genetic counselor) for clinical testing. However, as noted by the genetics clinicians that we interviewed, insurance coverage of exome and genome sequencing has improved over the past several years, particularly for families with public insurance. This, in turn, impacts the research referral pipeline. One clinician noted that they would generally refer patients to research only if they had prior negative testing and genome sequencing was not covered by insurance, which is becoming less frequent.

> “…But as genome is becoming more accepted… from a clinical ordering standpoint, we aren’t having to look to research opportunities to facilitate this genetic testing for our patients anymore, or as often” *Clinician 12, White female, pediatric geneticist*

The benefit of genome sequencing via clinical pipelines relates to a more reliable product and turnaround time, and due to this difference, clinicians saw RGP as the option only after they have tried to send testing to a clinical lab.

> “If I have not exhausted insurance options, I will probably work with insurance first and foremost. The reason is turnaround time. You know, that’s one of the things that patients want so much, is to have a prompt turnaround time. And so that’s what the commercial world is for.” *Clinician 16, White female, pediatric genetic counselor*

Given the difference in timelines between return of research results and clinical results, reliance on research studies to provide genomic services to families facing financial barriers to clinical access has the potential to worsen inequities. This has important implications for the consideration of the impact of research and clinical testing availability on equitable access to care. One clinician noted that “the care that a patient gets unfortunately is dictated, in some way, by their insurance coverage”, although substituting research genomic sequencing can “even that playing field a little bit. But it’s not like apples to apples, when it comes to those things.” (Clinician 9, White female, neurology genetic counselor). Clinicians also noted that relegating minoritized populations to research pathways for access to genetic diagnostic technology that is otherwise clinically available can worsen inequities.

> “… you know, what if it is something that there’s really important treatment or even potentially therapy for? And you know, they’re not getting a result for a year? …Whereas, if we had been able to do a test at a commercial lab and had that result back in six weeks, their management would have been handled more quickly. I think that would be the one main thing if an underserved population can’t achieve genetic testing in a commercial lab.” *Clinician 16, White female, pediatric genetic counselor*

#### Subtheme 2: Research may improve the clinical care of others

Although most families identified a potential clinical impact for their own family as a major motivator for research participation, many also expressed an altruistic aspect as well by contributing to scientific advances that could benefit the broader rare disease community. One participant noted that “…you might find out some things about my son that might be beneficial to other children that are like him also.” (Participant 1, Black or African American female, parent, sample pending). Another stated that “…if you don’t participate, well, it’s also more difficult for science to ever reach a solution.” (Participant 21, Hispanic female, Spanish-speaking parent, solved). Clinicians, too, felt a moral imperative: “this lofty goal of contributing to the cause … it’s to help patients with the idea that I’m also helping the larger cause.” (Clinician 7, White female, geneticist). In particular, participants felt a desire to spare others what they themselves had gone through, particularly those who felt that clinical benefit may be out of reach:

> “That’s the most important part, if they could help me or other people, because I’m so old that I think I’ve done all that I can do. But if they can help other people, younger people, or anyone else, you know, and I figure that’ll be the best part about it. If what I’m going through can help someone else, it’s more important to me than it is helping me, you know. I figure, if I got to go through what I’m going through to help someone else, I will do it. I think that’s the best part about this study, that it’s the idea that it may help someone else.” *Participant 6, Black or African American male, proband, analysis in progress*

Improving clinical care for future patients was also particularly meaningful for those who identify within minoritized racial or ethnic populations and the clinicians who care for these families due to the need to improve diversity in genomics. For example, as a parent of an affected child whose racial and ethnic identity has been historically underrepresented in genomic research, one participant expressed how this influenced their choice to enroll in research:

> “You know, another thing, too…is that there’s not a lot of African American genes in the database, because a lot of African Americans don’t put forth their genes into the—their DNA into the database. So it’s a constant struggle to sometimes get these conditions, figure out different genetic conditions that could happen, because there’s just not enough information. And they need information from other cultures… And so, to know that my son’s DNA will be added to the database, and will maybe help other people find a diagnosis sooner than what I had to do—Like I always think about that as well. Like how could this help other people, so that they don’t have to go through what I went through?” *Participant 7, White female, parent, analysis in progress*

One clinician who had the experience of having their patient diagnosed via participation in RGP reflected on the global impact of rare disease genomic research, noting that the finding of a causal variant in one person can facilitate the diagnosis of others and thus “… every person that’s involved in clinical research is hopefully helping themselves and is hopefully helping the community” (Clinician 13, White female, pediatric genetic counselor.). This potential for clinical benefit to oneself or others outweighed potential risks for multiple participants.

#### Subtheme 3: Research as a component of clinical care

The potential clinical impact of research participation directly relates to its incorporation into clinical care, both to overcome cost barriers as previously discussed and also to continue the diagnostic odyssey following non-diagnostic clinical testing by applying techniques that are not accessible to clinicians yet. Research has the potential to address access barriers, as one participant noted that they have trouble even accessing a genetic subspecialist, stating that “…it is difficult to convince a primary care doctor to refer you to a special geneticist. Trying to see a specialist called a geneticist is like pulling teeth in a lot of plans and a lot of care facilities” (Participant 5, White female, proband, analysis in progress). However, the implications of research studies as a substitute for clinical care must be considered, particularly as participants expressed the vulnerability of symptomatic individuals when being approached for research: “If there’s even a slight chance that you’ll get an answer when you’re sick, you’re going to take it.” (Participant 9, White female, proband, analysis in progress). Clinicians predominantly reported using referral to a research study as a way to potentially end the diagnostic odyssey:

> “But I would say, it’s sort of in concert with the work that we do. Like, clinical genetic testing has not been valuable for you. So, therefore now, we’re reaching into the research world where we’re continuing to learn more and continuing to dig into these things that we just don’t know yet.” *Clinician 13, White female, pediatric genetic counselor*

Clinicians and participants noted that genomic research studies have the capability to offer “testing that we clinically can’t get” (Clinician 8, Asian female, geneticist) because genomic technology evolves faster than the development of commercial labs to offer such testing. In this way, clinicians can see the research study as an extension of clinical care: “like every time someone has a negative exome, we just put a paragraph at the end of our note that gives them your email, website, and phone number” (Clinician 3, White female, geneticist). Participants, too, perceived this benefit of research.

Regarding the complex interplay between research and clinical care, clinicians also identified certain scenarios where they might recommend research specifically to fill a clinical gap - such as when a clinical test does not yet exist. Another reason for the use of research to achieve clinical goals is the ability to leverage novel genomic interpretation pipelines or identify novel disease-gene associations.

> “For example, the other day, I had a family with brain arteriovenous malformation… And we did a gene panel which came back negative. Usually, we are done, because for this phenotype…. we did all of the gene testing that we know already. But I was thinking about Rare Genomes Project, because you might be able to identify variant in genes that have not really been described formally.” *Clinician 6, Asian male, geneticist*

However, the distinction between clinical care and research was also noted by clinicians, sparking caution while at the same time holding potential to improve equity in genomics.

> “After all, it’s a research study and I have to be very cautious about using it for clinical care. But I do tell them that this is one way to leave no stone unturned to make a diagnosis, if there is a diagnosis to be made.” *Clinician 10, Asian male, physician in renal genetics*

Participants may not initially have a realistic view of the likelihood of diagnosis and resultant clinical benefit through his process. Discussion of the potential for clinical impact while also acknowledging the ultimate goal of research to generate general scientific knowledge allows for balanced counseling on research participation. Thus, this theme illustrates that the clinical impact of research participation relates to equity in access to genomic advances both at the personal/familial and community level. However, given the differences in clinical and research workflows, care must be taken to avoid exacerbating inequities.

#### Theme 2: Research participation functions as a rare disease community resource, offering psychosocial benefit and access to emerging science

This theme encompasses the idea of research participation as “an end in itself”, with benefits to the family even if a diagnosis or cure is not found and separate from altruistic reward.

Participants and clinicians described the benefit of continuing to search for an answer (subtheme 1) which inherently validates parental or personal concerns regarding their symptoms. Another subtheme is the idea of generating community via research participation (subtheme 2).

#### Subtheme 1: Psychosocial benefit - “the wheels are still turning”

First and foremost, many participants and clinicians felt that a genetic diagnosis would represent a validation of medical concerns that had been raised and, sometimes, ignored. One participant stated that they were searching for such validation via potential diagnosis through the research study:

> “I was hoping to be heard, to be believed, to be accepted, and to finally have a resource that may get me somewhere with my care… to finally have a resource to improve my care, and communicate with physicians in the future.” *Participant 5, White female, proband, analysis in progress*

In addition, research participation was noted to offer mental relief from searching for an explanation for the rare disease in their family, whereby clinicians noted that the search itself can be validating, in the feeling that “the wheels are still turning… there are people continuously looking at this data to try to figure it out” (Clinician 11, White female, neuromuscular genetic counselor), regardless of whether an answer is ultimately found. Participants also expressed deriving benefit from knowing that the research never stops looking:

> “And there was one other thing, that even if they can’t find something, which was very interesting and cool, is that if they don’t come up with the resolution, they don’t kind of cancel you out. They work longer with you and they kind of, I don’t know how to word it, but kind of like keep you in mind for future testing. So it seemed like it was like an ongoing thing and that they didn’t give up on you, which was really cool.” *Participant 22, American Indian or Alaska Native and Black or African American, female, proband, sample not received*

Given these psychosocial benefits which may be present even in the absence of a diagnosis, many participants expressed that they simply saw no particular downside to research participation, that it was a “win-win” situation (Participant 1, Black or African American female, parent, sample pending).

#### Subtheme 2: Finding community in research participation

Both clinicians and participants acknowledged a feeling of isolation due to having a rare condition. Some participants expressed a desire to find a community through diagnosis, while others sought a sense of community in joining other parents who are pushing the clinical boundaries to help their children. This idea of supporting “the cause” was expressed by one participant who felt that this benefit negates potential downsides of research participation:

> “I’d like to join forces with other people who are dealing with the same conditions as my daughter, and with rare diseases, right? Because you can learn or you can help in another way, and another person can help us in one way or another, you know. We can help each other, you know, we can try to help..” *Participant 18, Hispanic female, Spanish and English-speaking parent, analysis in progress*

However, participants also recognized that not everyone may perceive a benefit from knowing the answer, even within a family. As one participant noted, she and her partner differed on their desire to find a diagnosis for their child: “culturally, like, just the way I am, I want answers. And with him, I think he’d be okay with not.” (Participant 7, White female, parent, analysis in progress). Another participant also expressed this divergent viewpoint within a parenting couple, which was informed by perception of disability:

> “I think that was another thing that my husband didn’t really speak on, but was concerned about. He doesn’t really—I think that he didn’t want [CHILD] to view herself as disabled, and doesn’t want to have that in the back of his mind that she has a disability, or she’s not capable of something because of having a disability. So I think that was a lot of his apprehension as well.” *Participant 12, White female, parent, lost to follow up*

In this sense, the connection to a community identity that may be centered in disability was not always desired and may negatively impact research participation.

#### Theme 3: Flexibility in research processes allows for increased access for diverse participants

Theme 3 reflects participant and clinician viewpoints on research workflows and accessibility for underserved populations. Participants and clinicians commonly shared that having an adaptable research process reduces structural barriers to study engagement. Both also expressed that broader participation has been enabled through the involvement of trusted clinicians (subtheme 1) and logistical flexibility that includes personalized support from the study team (subtheme 2).

#### Subtheme 1: Partnership with trusted clinicians

Clinician referrals, clinician application support, and clinician endorsement of the RGP has helped engage families, especially families who commonly experience barriers in healthcare. Generally, participants and clinicians have described the overall referral and application process as accessible, which subsequently encouraged future referrals from clinicians. Having different referral pathways has allowed clinicians to be as involved as needed, to support their patient population or also to navigate their competing clinical demands.

Most participants in the RGP self-applied to the study through the study website. In addition to being an accessible overview for self-referring families, this website serves as a good reference point for clinicians to give their patients who may be self-motivated to apply but have not heard of the RGP previously. Being familiar with study eligibility criteria, some clinicians offer additional support for patient-initiated applications by either letting their patients know what details to include or helping their patients fill out the online application through the study website. One clinician recognized that their patients were not completing the online application and coached them through the process, because “there’s a bit of follow through that’s sometimes necessary” (Clinician 15, Asian female, pediatric genetic counselor).

Separate from the RGP online application, other clinicians have completed a clinician referral form, on behalf of their patients who they “suspect will have barriers to their care” (Clinician 5, White female, geneticist). When describing completing a clinician referral form, clinicians appreciated the ease and brevity of the form. Notably, participants also appreciated the clinician support, as showcased by a participant who credited their study enrollment to their referring clinician:

> “You know, I think if it wasn’t for my doctor putting in all the information, I don’t know if that was something that I would have to do. And I think that would have been difficult. But in my case, she did her part and it made that transition really smooth for me to kind of do what I had to do. And she did all the medical stuff, which, you know, your typical person isn’t going to understand all the medical terminology. So I think that that makes it easy when the doctor is doing the medical stuff.” *Participant 20, Hispanic female, parent, solved*

Another benefit to working with clinicians was the opportunity to set expectations early on, particularly related to the longer turnaround time for research results, if any are found. This subtheme thus reveals the importance of partnership with clinicians in designing inclusive research practices.

#### Subtheme 2: Remote design, flexible logistical options, and individualized support

The remote and no-cost study design of the RGP makes study participation attainable for those where travel, time, and financial resources may be a barrier. One clinician described this study as a realistic recommendation for motivated families with non-diagnostic comprehensive clinical genetic testing or with insurance barriers:

> “Because again, I don’t like to recommend things to people that it’s really not realistic for them to do. Like they don’t have the money to miss a day from work… So I feel like, with [the RGP], it doesn’t seem like there’s a big commitment on the part of the patient, other than providing their medical information and a blood sample. And that’s more like the speed of what many of our patients would be able to handle.” *Clinician 3, White female, geneticist*

Flexible timing, such as asynchronous options and varied scheduling options, allows individuals to participate in ways that align with their availability, especially for those with caregiving responsibilities or work commitments. As explained by Participant 4 (Asian female, parent, primary language - Urdu, analysis in progress), the remote study design was favorable for busy parents who are already managing complex medical care for their child. Language flexibility was also important, where having Spanish-speaking study staff and interpretative services increased participant understanding, encouraged rapport, and established a level of comfort among study enrollees, as expressed by a Spanish-speaking parent who stated: “I can get by in English, but when it comes to money and genetics, please speak in my native language.” (Participant 14, Hispanic female, sample pending). Furthermore, another Spanish-speaking parent (Participant 21, Hispanic female, solved) highlighted a preference for direct communication with Spanish-speaking RGP staff over the use of interpretative services, noting a concern that external interpreters might occasionally misinterpret or alter the intended meaning of the conversation.

Through virtual consent sessions with study staff and sometimes through referring clinicians, RGP study staff can identify prospective participants who may need individualized support, such as mobile phlebotomy. One geneticist stated that referrals are made easy because “the research team would do the direct contact with the families if they were not willing or able—mostly able—to do the initial reach-out themselves” (Clinician 5, White female, geneticist). Another clinician also appreciated when the study staff could proactively reach out to their patients, when necessary, stating “having somebody be able to proactively reach out to the family has been like the number one barrier removed” (Clinician 13, White female, pediatric genetic counselor).

While the remote study design was feasible for most participants, it does not work for all. When discussing what makes study referral difficult within their patient population, Clinician 13 shared “the biggest thing with the families [experiencing housing instability] that we have [is they] just don’t have reliable access to technology.” Offering multiple ways to connect, such as phone, email, and postal mail, provides accessibility to those who may have limited access to the internet or to those who may not be digitally adept. For example, one participant (Participant 17, Black or African American female, proband, analysis in progress) mentioned that they were able to electronically sign the consent form, but their mother preferred the consent form be sent through postal mail. Other participants stated that they do not have regular access to email and communicating via this modality resulted in delays in enrollment. To complement the digital resources, clinicians have recommended the distribution of physical study flyers to their clinics—a resource currently offered by RGP—serving as an additional reference alongside the patient-friendly website. Thus, again, access was noted to be both facilitated and blocked by advancing technology.

Finally, collaborating with a laboratory network that has various locations nationwide facilitated no-cost sample collection, permitting enrollees to schedule phlebotomy at their preferred time. However, for individuals in rural areas lacking nearby lab locations or those with limited physical abilities, study accessibility was enhanced through mobile phlebotomy services or the option to utilize a known clinician’s office to have the sample collection. Since the contracted laboratory operating hours conflicted with parental employment commitments, mobile phlebotomy services facilitated successful sample collection for Participant 19 (Hispanic female, Spanish-speaking parent, analysis in progress) and their family.

#### Theme 4: Longitudinal engagement is strengthened through relational continuity with participants and their clinical team

Beyond procedural flexibility, participants highlighted the importance of responsive and understanding interactions with study staff (subtheme 1). These interactions built trust and encouraged continued participation, reinforcing access not just as entry, but as sustained engagement. Additionally, for clinicians, continued engagement is noted to be conducive to their affinity for referring eligible participants to our research study (subtheme 2).

#### Subtheme 1: Positive interactions between participants and study staff

Eligible applicants for RGP engage with study staff prior to enrollment during a video or phone call informed consent session, which lasts roughly 30-45 minutes. Throughout this meeting, individuals are provided with a comprehensive summary of the study goals and process. This is an opportunity for prospective participants to ask a member of the study team questions about our study before enrollment. Participant 5 (White female, proband, analysis in progress) described the study team’s “demeanor, professionalism, friendliness, and patient-focused approach” during that initial communication as a “driving force” in their decision to participate in this research study. Study participants often noted that meaningful human connection with researchers was important, specifically valuing a compassionate listening ear and the opportunity to receive personalized clarification from a person rather than automated messaging:

> “…People who care makes it easy to join. People who just, you know, show that they are working towards a common goal, which is what has been displayed to me. And so far, that’s what has prompted me to just want to be a part of it… They want people who are willing to be compassionate and kind, and are willing to just be willing to listen. And that’s a big thing, is listening and being kind.” *Participant 2, Asian female, proband, analysis in progress*

For Participant 5, the virtual consent session occurred at a time when they felt overwhelmed and frustrated by their diagnostic odyssey; while experiencing a nosebleed, a clinical symptom of their condition, during the call, they found that the team’s response provided the necessary support:

> “The thing that I needed, that they gave, was empathy. Needed it, got it. And it was so badly needed at that point, that the girl who took my call couldn’t have handled it better, or couldn’t have provided what I needed better. She immediately led with empathy, led with understanding, and said, ‘I cannot imagine what this has been like for you.’ And then I had the video interview. And I’m bleeding. I have a nose bleed during that video interview. And that person expressed empathy. That’s what I needed, and that’s what I got.” *Participant 5, White female, proband, analysis in progress*

Following their study enrollment, participants typically maintained communication with study staff through email or telephone. These touchpoints were frequently utilized to coordinate the collection of relevant medical records, relay changes in clinical presentation, or inquire about their current progress within the research pipeline.

#### Subtheme 2: Maintaining relationships with clinicians

Sustained longitudinal engagement with referring clinicians during the RGP process is supported by regular and consistent communication, a process that clinicians highlighted as a necessary area for continued improvement during interviews. These professional relationships are instrumental in the identification of eligible participants for the study. For a family navigating the disappointment of negative clinical genetic testing, Participant 22 (American Indian, or Alaska Native and Black or African American female, proband, sample pending) reflected that their clinician’s description of the project offered “hope [that] it’s not over,” reinforcing the possibility of attaining a definitive genetic diagnosis.

Clinician engagement generally occurred at two primary milestones: the initial study referral and the return of a genetic finding from our study. Outside of these touchpoints, some clinicians occasionally inquire about the status of their patient in our research pipeline. While most appreciated the hands-off nature of the study’s logistical coordination, Clinician 12 (White female, pediatric genetic counselor) noted a preference for a more integrated role, suggesting a desire for involvement and consistent notifications throughout each phase of the process. For others, the experience of delivering results has served as a motivator for future patient referrals. Beyond the delivery of genetic findings, Clinician 4 (American Indian or Alaska Native & White male, neurologist) valued the supplementary clinical guidance provided during the research result return process as well. Clinicians whose referred patients have yet to attain a genetic result through our research study expressed that this lack of diagnostic results can negatively impact their affinity for future referrals, suggesting that the absence of findings has diminished their perceived utility of the study. One clinician characterized the lack of consistent feedback as a “black hole,” highlighting a significant disconnect in communication. To remedy this, some proposed that the study team initiate proactive status updates regarding their referred patient population.

Considering the themes that arose from the clinician and participant narratives, we have synthesized both recommendations for future clinical and research practice as well as areas in need of further research (**Table 3**):

**Table 3.** Considerations for the future.

| Table 3. Considerations for the future |  |  |
| --- | --- | --- |
| Theme | Recommendations | Future Studies |
| Theme 1: Rare disease genomic research studies are intertwined with clinical care and hold potential overcome access barriers in the clinic, with subsequent equity implications. | Distinguish between clinical and research genomics, ensuring clinicians can manage expectations about diagnostic yield without diminishing the potential value of the study to the family and to the broader community | Further quantify the psychosocial benefit participants gain from research participation during their diagnostic odyssey |
| Theme 2: Research participation functions as a rare disease community resource, offering psychosocial benefit and access to emerging science. | Point families towards clinical testing options first if they/their clinicians were not aware of this possibility, reserving research opportunities for after clinically available testing options have been |  |
|  | exhausted, either due to access or other barriers or negative testing result.<br>Explore low-burden ways to foster connection between participants, such as newsletters that share collective study progress rather than just individual results |  |
| Theme 3: Flexibility in research processes allows for increased access for diverse participants. | Shape research logistics to adapt to the individual participant<br>Language-concordant support from study staff directly when possible and with interpreters and translated documents when not<br>Prioritize opportunities for human connection between staff and participants<br>Develop a standardized communication cadence that provides referring clinicians with proactive status updates on their referred patients<br>Establish relationships with clinicians who serve historically marginalized populations | Gather perspectives from participants who declined research participation or were lost-to-follow-up<br>Evaluate success of partnership with trusted non-genetics clinicians |
| Theme 4: Longitudinal engagement is strengthened through relational continuity with participants and their clinical team. |  |  |

**Table 4.**
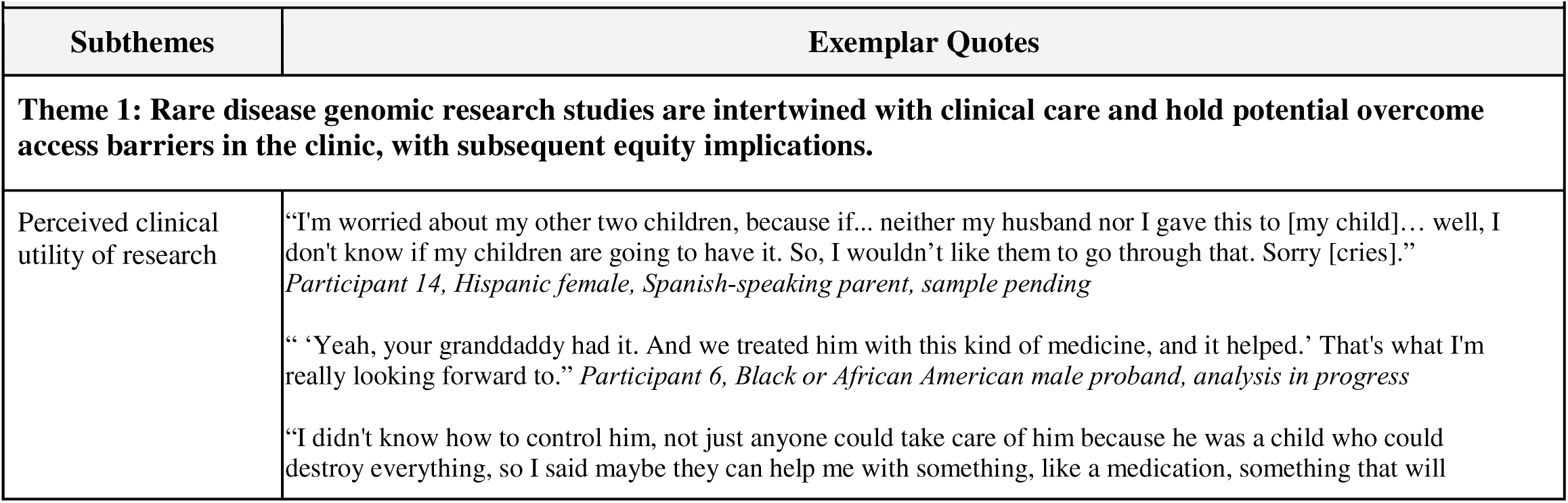
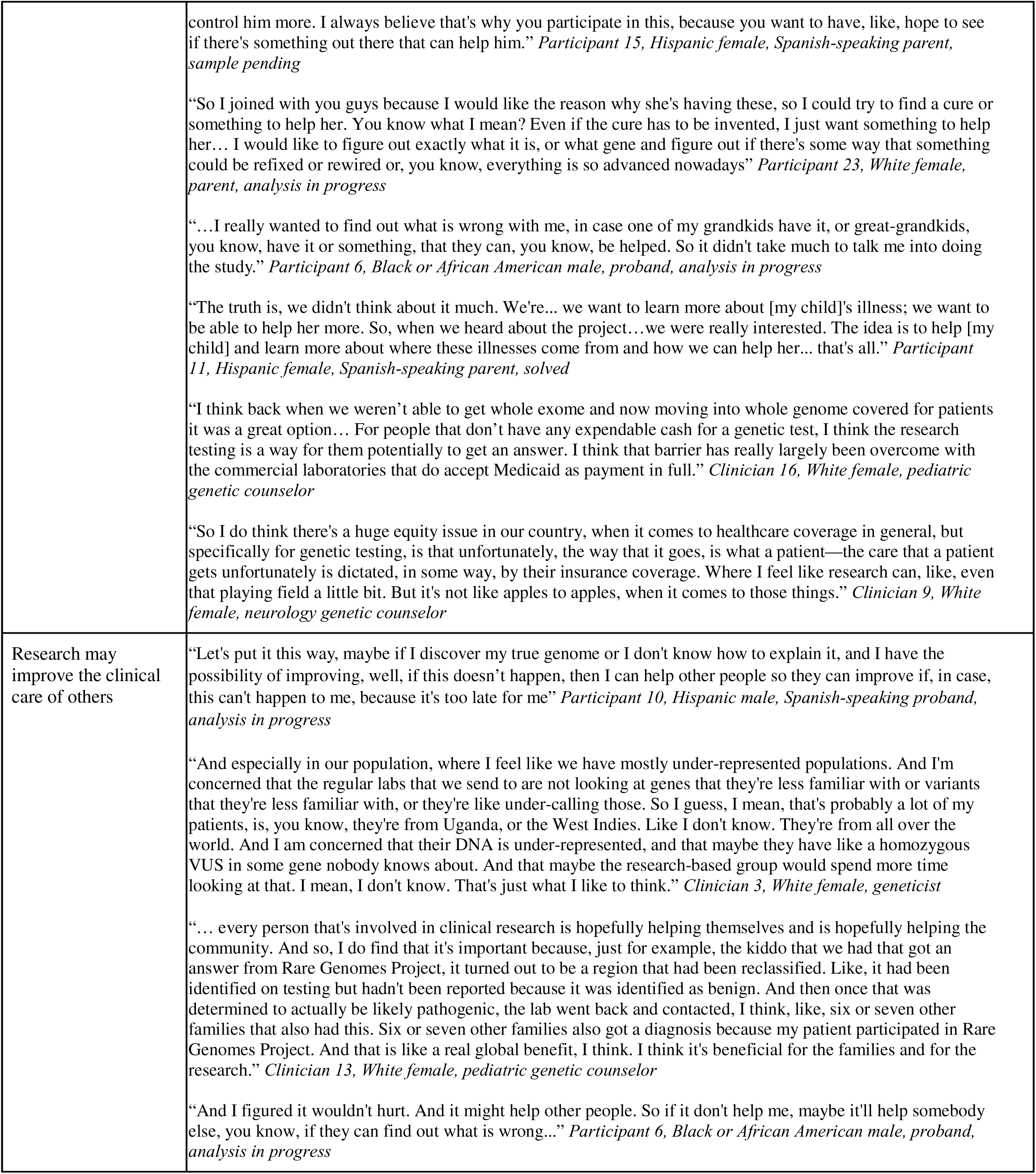
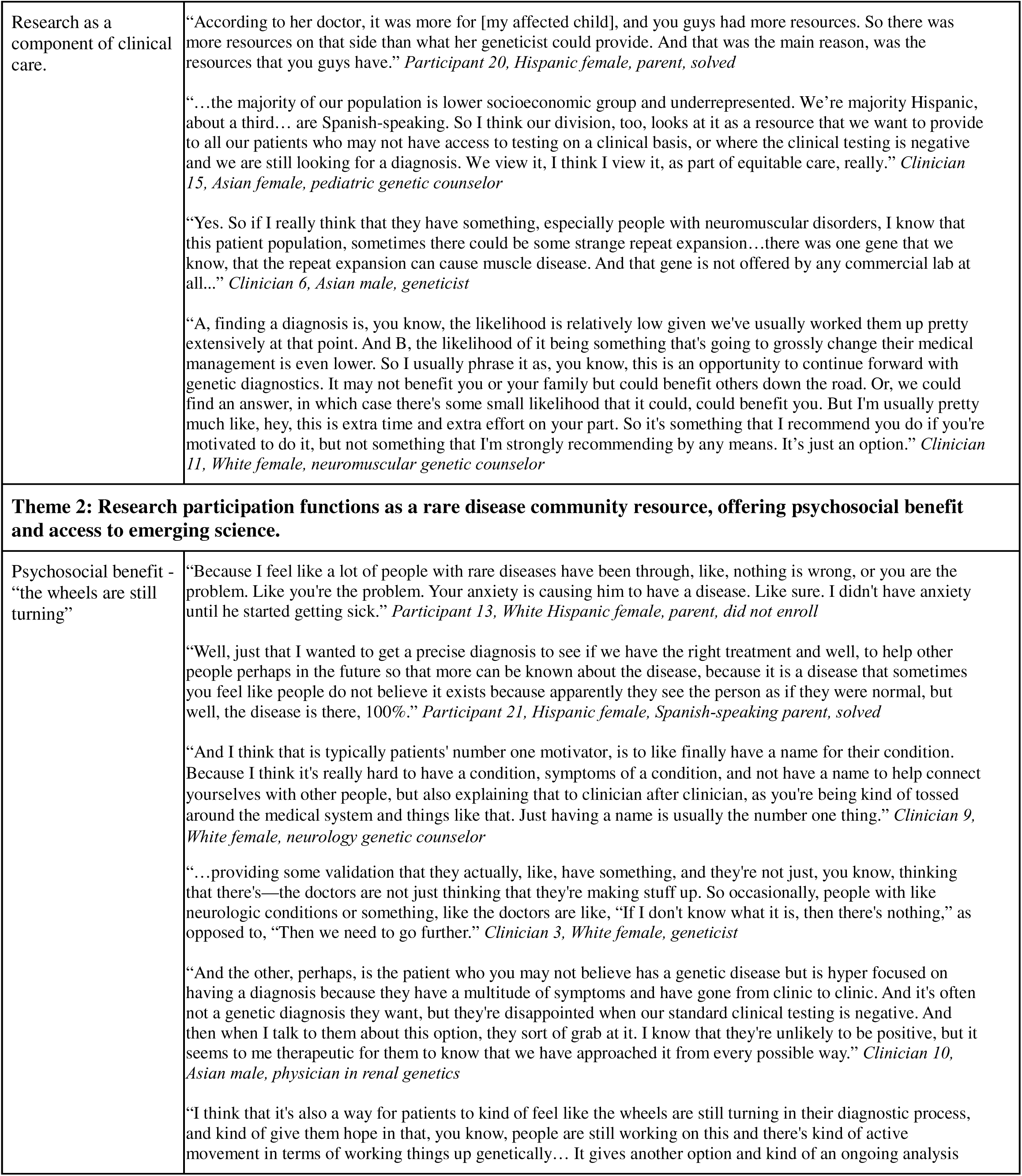
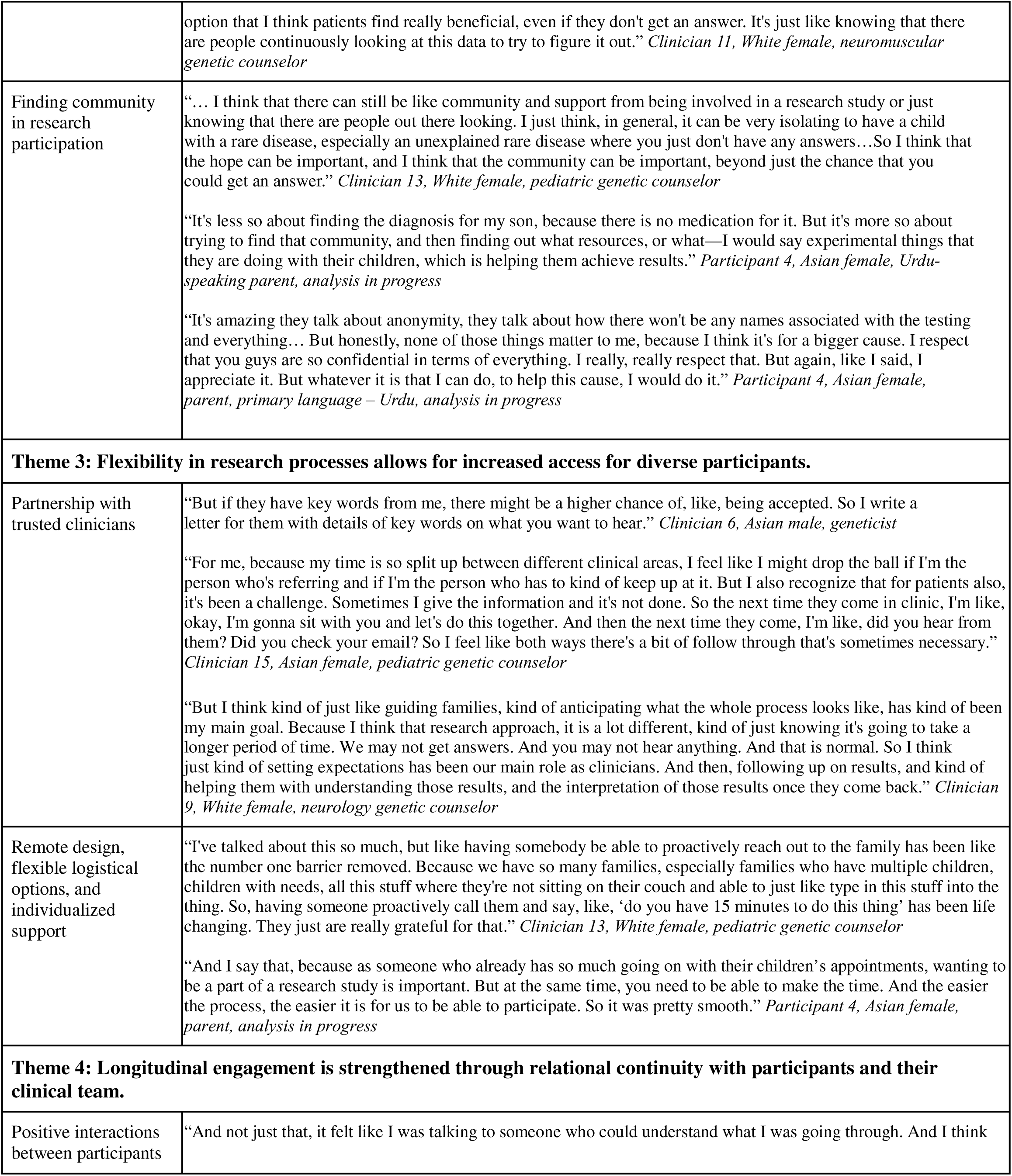
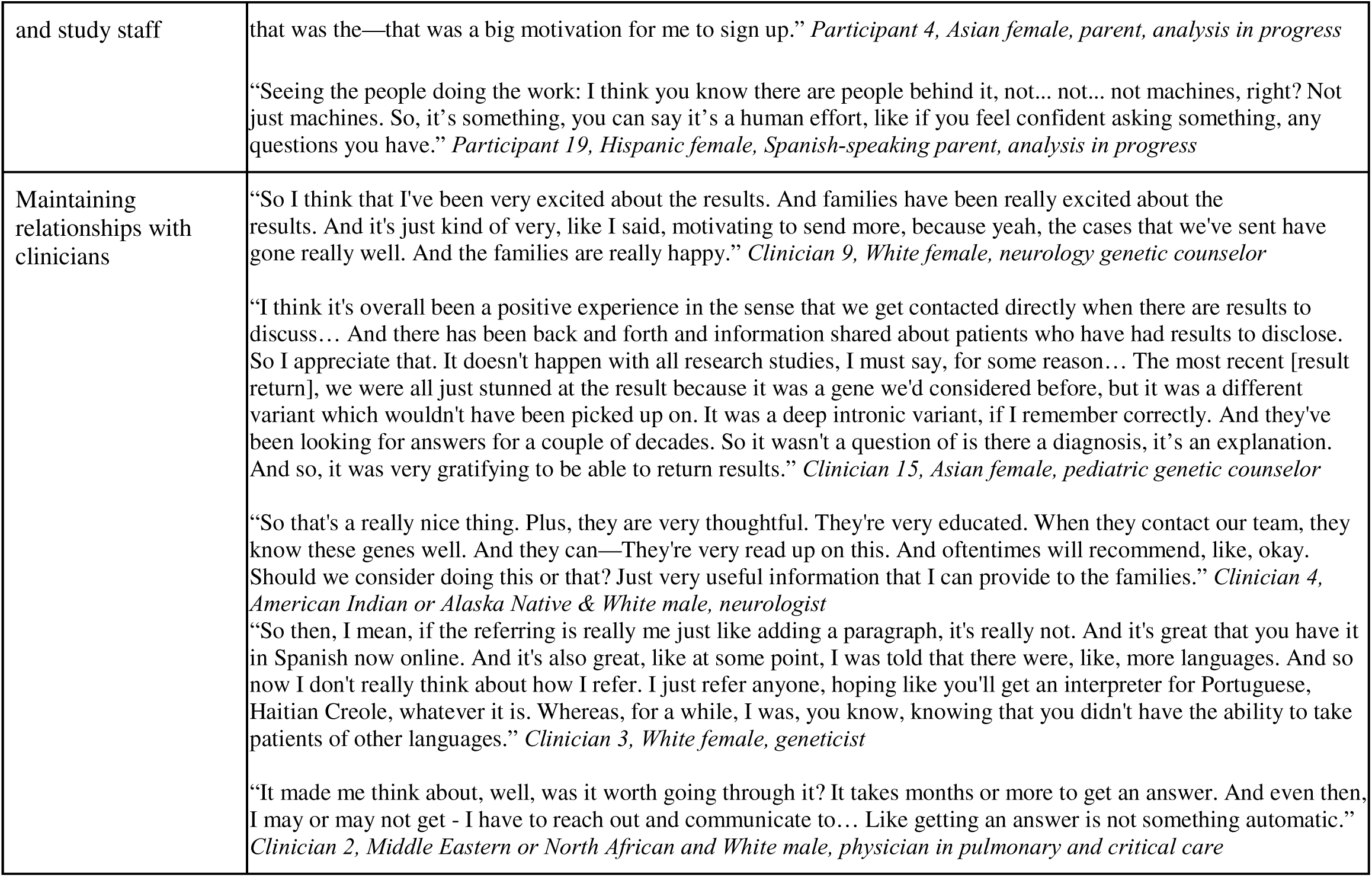
Exemplar Quotes.

## Discussion

We present a qualitative analysis of participant and clinician perspectives on rare disease genomic research participation, with a focus on access equity for populations facing historical barriers to research or clinical genomics. One strength of our study is the inclusion of diverse participants in order to gain additional insight into the perceived value of genomic investigation for rare diseases, where most prior studies reporting on utility of genomics include white women with relatively high educational and income status^21^. Woven throughout these narratives is the interplay between research and clinical genomics, particularly pertaining to the use of research as a means to access a genetic diagnosis if such clinical genetics services or genomic sequencing are not otherwise freely available. It is worth noting that although the Rare Genomes Project returns results to enrolled families when a potentially diagnostic genetic finding is identified, this practice varies across the research protocols of other genomic studies. Both participants and clinicians recognized the ability for research to provide access to genomic technologies not available routinely in clinical practice, particularly for historically underserved populations facing barriers to access to genetics subspecialty clinics or access to sequencing due to cost and insurance coverage. However, both also acknowledged the potential dangers of research filling a clinical need. This has equity implications as well, as those who have no access to clinical testing may be more enthusiastic about joining a research study, where the testing itself is a form of enticement. At the same time, multiple participants also reported feeling a human connection or empathy from the research staff that they had not found in their clinical journey, illustrating another pathway by which research processes may inform clinical practice.

### Lessons for the clinic

Although not intended to serve as a replacement for clinical care, experience with genomic research may inform clinical workflows as technologies such as ES and GS are now becoming more accessible for both genetics and non-genetics clinicians. As clinical GS becomes more accessible, the workflow enablers that were identified in these interviews may cross over to clinical genomics, rather than solidifying research as the path to GS for vulnerable populations. Previously reported barriers to clinical and research genomic sequencing include lack of time and lack of knowledge about genetic research^21,22^. One unique feature of our study was the focus on concrete logistical steps that block or facilitate access. We identified that rather than supporting the participant to fit the process, successful genomic research projects help shape the study processes to fit the participant. By maintaining an adaptable research framework, RGP actively dismantles preconceived notions regarding accessibility. Feedback from our enrollees underscores that logistical preferences are highly individualized; consequently, a process that facilitates engagement for one family may present unforeseen challenges for another. In the clinic, this suggests that a focus entirely on telemedicine and web-based support, for example, for GS may not improve access for all. Ideal workflows would maintain flexibility and adaptability. Finally, as in prior studies, the importance of language-concordant support was repeatedly expressed among those whose primary language was not English^21^.

Another important facet of our interviews was the human connection that participants felt with the research team. While some participants were primarily motivated by their search for a genetic diagnosis, others emphasized that their decision to enroll was encouraged by the personalized guidance and support received from both their clinical team and the research staff. While participants discussed mistrust of the medical system, in some ways they noted a stronger connection to the research team because they took the time to listen to the person’s medical history and validated their concerns. Even though the RGP process is largely remote, the availability of genuine human connection between research personnel and prospective participants remains a critical factor, frequently serving as the primary catalyst for a family’s decision to engage with our genomic study. Thus, although electronic consent is widely regarded as a streamlined and accessible pathway for enrollment, a human-less process may not be superior to face-to-face methods: particularly for populations that have historically been mistreated by medical researchers and clinicians.

### Lessons for future research studies

The research participants that we interviewed also offered insight into their motivations for participating in genomic research. They highlighted that the reasons for research participation are distinct from motivators in the clinical setting, such as impact on care. Gaining insight into the core elements that drive participation in genomic studies, from both the clinician and participant viewpoints, may enhance recruitment approaches designed to bolster the inclusion of historically marginalized families within the rare disease community. Prior research from the Undiagnosed Diseases Network described that their participants, particularly Hispanic participants, endorsed relying on trusted clinicians for access to genomic research. For populations with less trust of the medical system (in their study, Asian or white families), higher social capital permitted continued access to genomic research despite mistrust^22^. We also found that endorsement from trusted clinicians facilitated access to the RGP from our participants, particularly for families with potential language barriers to clinical care. Some participants that we interviewed also endorsed experience with medical gaslighting or mistrust of the healthcare system, and the separation of the research team from this system in some ways empowered participants to enroll.

Our study is novel because of the unique perspectives of referring clinicians. Prior rare disease genomic research studies describe recruitment pathways supported by clinical geneticist and genetic counselor referrals^15, 23^. However, this work does not elucidate the reasons for such referrals. By incorporating the referring clinicians’ perspectives, our qualitative analysis contributes to the field by identifying specific clinician motivators that drive patient engagement with research. We found that clinicians have different reasons for referring families, and this may inform family mindsets and views on participation.

Overall, the motivators and experiences of the diverse participants that we interviewed highlight the importance of providing equitable access to genomic sequencing for rare disease diagnosis while working responsibly within healthcare systems. Interestingly, in terms of the mindset of people with rare diseases who are interested in genomic sequencing, our participants did not regularly reference cultural norms when thinking about rare disease diagnosis or research participation, in contrast to prior research with Hispanic and Asian families^19^. Instead, they expressed viewpoints were often aligned with a recent qualitative analysis of the experiences of people with disabilities participating in the UDN, where flexible procedures and supportive staff were highly valued^23^. Lessons learned from genomic research studies may inform clinical practice as ES and GS are more routinely covered by insurance and offered by health systems. In addition, by integrating genomic study opportunities into standard clinical workflows, clinicians can effectively treat enrollment as an extension of comprehensive patient care. Collaborative partnerships with these clinicians serve as a successful mechanism for identifying eligible families who may benefit from research participation but are unaware of such opportunities and/or face obstacles to access. Given that underserved populations experience disparities and delays in accessing clinical genetics services^24,25^, future work should focus on the success of collaborative partnerships with non-genetics clinicians to broaden clinical and research access within these communities. Ideally, even as artificial intelligence, chatbots, and digital tools are increasingly becoming prevalent in medical care, particularly genomics, people with rare diseases will find the human empathy and support that they desire in the clinical space.

## Data Availability

Data produced are available contingent upon data use agreement.

